# Effect of kettlebell training on bone mineral density in two older adults with osteoporosis: a multiple-case study from the BELL trial

**DOI:** 10.1101/2021.08.15.21261771

**Authors:** Neil J. Meigh, Justin W.L. Keogh, Wayne Hing

## Abstract

The purpose of this explanatory retrospective case study was to report clinically significant increases in bone mineral density in a female and a male over 70 years of age with osteoporosis, following 16 weeks of hardstyle kettlebell training. Both case subjects were insufficiently active prior to participating in the BELL trial. Subjects trained five days a week accruing a large training load volume (calculated as kettlebell mass multiplied by repetitions performed) during structured group-based classes (74,872 kg and 110,132 kg, respectively). Regional dual-energy X-ray absorptiometry was used to assess BMD at the hip and lumbar spine. Increases in BMD of 12.7% and 5.9% at the femoral neck and lumbar spine (L2-L4) respectively were observed for the female, and 2.5% and 6.0% respectively for the male. Magnitude of change in BMD (g/cm^2^) at the lumbar spine was 2.0 and 1.9 times larger than the least significant change for the female and male respectively, and sufficient to advance the female subjects’ status from osteoporosis to osteopenia. Although these results do not show a definitive causal relationship between kettlebell training and increased BMD, further investigation of the effects of kettlebell training on BMD in older adults with osteoporosis and osteopenia is warranted.

## 1. Introduction

This study describes clinically significant increases in bone mineral density (BMD) in two older adults with osteoporosis who participated in the BELL trial ^(1)^, which involved group and home-based exercise with kettlebells performed five days a week for 16 weeks. Osteoporosis is a systemic skeletal condition associated with ageing, characterised by very low BMD. Osteoporosis is diagnosed using dual-energy X-ray absorptiometry (DXA) from a T-score more than 2.5 standard deviations (SD) below that of sex-matched young adults, whereas osteopenia (pre-osteoporosis) is diagnosed when the T-score is between 1 and 2.5 SDs below sex-matched young adults ^(2)^.

Bone metabolism deteriorates with advancing age. Rates of bone loss increase from 0.6% to 1.1% annually, in postmenopausal women 60-69 and 70-79 years, respectively ^(3)^, which can be exacerbated by inactivity, disuse, trauma, and medical conditions such as diabetes. Currently, approximately two thirds of Australians over 50 years of age have low bone mass ^(4)^, with 500 daily osteoporotic fractures projected to occur in Australia by 2022 ^(5)^. More than one in three women and one in five men are likely to sustain at least one fragility fracture in their lifetime ^(6, 7)^ with the cumulative risk of fracture for postmenopausal women as high as 60% ^(8)^. More than 50% of older adults who fracture a hip are no longer able to live independently and almost one in three can die within 12 months of the fracture ^(6, 9)^.

Between 2007 and 2017, the direct costs of osteoporosis are estimated to have tripled ^(10)^. Hip fractures account for the largest proportion of this increase (43%), with 74% of the fractures occurring in people over 70 years of age ^(10)^, with direct and indirect costs of osteoporosis projected to be $3.84 billion by 2022 in Australia ^(5)^. In addition to the financial cost, the social burden is considerable, with fragility fractures, especially in the hip and spine, causing severe pain, disability, loss of independence, and decreased quality of life ^(7)^. Up to 70% of the variance in fracture from loading is explained by BMD ^(11)^ with each 1 SD reduction in BMD representing a 1.5 to 3-fold increase in risk of fracture ^(2, 6)^. Combined with other benefits of structured exercise, such as improved muscle function and balance, a 1-2% increase in BMD via an exercise prescription may reduce fracture risk by 5-10% ^(12)^ and reduce the incidence of fracture by up to 50% ^(13)^. Thus, research to determine optimal exercise prescription for improving BMD remains a high priority.

Exercise prescription and lifestyle modification are interventions for osteoporosis which aim to promote bone health ^(2)^. Fracture index is calculated from BMD T-score, so therapeutic interventions which positively influence T-score have the potential to reduce risk of fracture. Exercise may increase bone mass and bone strength, however effects from clinical trials appear to be site specific and provide only small clinically relevant improvements in BMD (<2%), and small absolute reductions in fracture risk, if at all ^(6, 8, 13, 14)^. Although the optimal type and dose of exercise is not definitive, current consensus is that a combination of high impact and progressive whole-body resistance training (HiPRT) is the most effective for maintaining bone mass (preventing bone loss) at the femoral neck (FN) and lumbar spine (LS) in older adults with low BMD ^(3, 4, 15)^. Walking, cycling, swimming and even impact and resistance exercise appear to be largely ineffective for increasing BMD when performed in isolation^(4, 16, 17)^.

Underscoring the prophylactic nature of exercise, clinical guidelines recommend that exercise should be lifelong and include regular weight-bearing and high impact activities to promote muscle strength, muscle power, and maintenance of bone mass and geometry ^(6, 8, 14, 18, 19)^. In the osteoporosis population however, adherence rates to exercise programs are generally poor, with a recent review suggesting that 50% of participants drop out within the first 6 months. ^(20)^. Results from the BELL trial, which maintained an adherence rate >90% for over 12 weeks, show that resistance training programs can be performed by older adults safely and independently with positive psychosocial affect, despite training five days a week and the intensity being “very hard” ^(1, 21)^.

Multiple studies, from single case reports ^(22)^ to large randomised controlled trials ^(23)^, have attempted to find the most effective types of exercise and program variables to improve BMD in older adults with osteoporosis. Results have been varied, from no significant difference after 12 months of training, to extraordinarily large increases many times greater than mean change reported in meta-analyses ^(14, 24)^. Novel methods of utilising resistance training equipment with older adults, such as kettlebells ^(25)^ and weighted vests ^(26)^, allow researchers to determine how changing program variables may influence the bone response to exercise. These investigations are valuable as exercise interventions thus far, appear to have been more effective at maintaining rather than increasing bone mass. The aim of this case study is to report the effects of a 16-week high volume hardstyle kettlebell training program on BMD and measures of healthy ageing in two adults with osteoporosis over 70 years of age.

## 2. Methods

### 2.1. Ethical approval

The study was approved by the Bond University Human Research Ethics Committee (NM03279) which was subsumed within a larger clinical trial. The BELL trial was pre-registered on the Australian New Zealand Clinical Trials Registry (ACTRN12619001177145). Permission to reproduce BMD reports was granted by both subjects. CARE (CAse REport) checklist. A copy of the CARE checklist is included in Appendix 26.

### 2.2. Case description

Pseudonyms are assigned for anonymity. Subject’s characteristics at baseline are presented in Table 1.

**Table 1.** Subject characteristics at baseline

|  | Sally | Peter |
| --- | --- | --- |
| Age (yrs) | 70-75 | 70-75 |
| Height (cm) | 159.0 | 176.8 |
| Body mass (kg) | 44.6 | 89.4 |
| BMI (kg/m<sup>2</sup>) | 17.4 | 28.6 |
| T-score FN | -2.3 | -3.11 |
| LS | -3.0 | -0.85 |
| DXA SMI (kg/m<sup>2</sup>) | 5.16 | 6.99 |

BMI; body mass index, FN; femoral neck, LS; lumbar spine, SMI; skeletal muscle index

**Case 1**: Sally was diagnosed with osteoporosis by DXA scan in November 2017. A second scan conducted in January 2020, revealed negligible change in bone mass in the two years immediately preceding the BELL trial intervention. Sally walked regularly and was otherwise in good physical health with no prior resistance training experience. Sally had not received any medications to treat her osteoporosis.

**Case 2**: Peter was diagnosed with osteoporosis by DXA scan in December 2019. A course of Denosumab (Prolia) was commenced in January 2020 less than a month before the BELL trial intervention commenced. Peter is a diabetic with peripheral lower limb neuropathy and mildly symptomatic knee arthritis. Peter worked part-time and his health was otherwise unremarkable, however, limited lower limb mobility and discomfort made floor transfers and lunging exercises challenging. Peter had some limited resistance training experience having previously engaged a personal trainer for approximately thirty, 30-minute sessions through 2016 and 2017.

### 2.3. DXA

Both subjects received regional DXA BMD scans before and after the training period, at the direction of their General Practitioner, and external to the BELL trial. Change in DXA-derived body composition was a secondary outcome of the BELL trial however, measures of whole-body composition are inadequate for accurately measuring regional BMD. Both subjects provided the BMD reports to the researchers with permission to reproduce results for publication. Peter’s pre- and post-intervention scans were conducted using the same Norland Machine. Sally’s scans were conducted on different machines (Hologic and Norland).

#### 2.3.1. Least significant change

Least significant change (LSC) for lumbar spine BMD was calculated according to the equation given by Nelson et al. ^(27)^ where LSC = 2.77 × precision [g/cm^2^], and precision = 0.00161 + 0.000364 × BMI [kg/m^2^].

### 2.4. Outcome measures

Measures of health-related physical fitness in the 6-month repeated measures BELL trial included: grip strength (primary), 6-min walk distance, resting heart rate, stair-climb, leg extensor strength, hip extensor strength, Sit-To-Stand, vertical jump, five-times floor transfer, 1RM deadlift, body composition (DXA), attendance, and adverse events. Due to COVID-19 close-contact restrictions at the time of testing, participants over 70 years of age, (which included Sally and Peter) were only permitted to perform three tests: grip strength, five-times floor transfer and 6-min walk, however, Peter did not complete the 6-minute walk test. Results from only these three outcomes are presented here. Full details of the outcome measures and trial results from mixed effects modelling have been published elsewhere ^(1)^.

#### 2.4.1. External training load (V-TL)

External training load volume was calculated as the product of [kettlebell weight (kg)] × [repetitions] performed within each session e.g., 8 kg × 100 swings = 800 kg. The overall training load for the 12-week intervention was calculated as the sum of the training loads for each individual session.

#### 2.4.2. Internal training load (sRPE)

Internal training load (arbitrary units, AUs) was calculated as the sum of self-reported [session rating of perceived exertion (sRPE)] × [training duration (mins)] e.g., sRPE 7 × 45 mins = 315 AUs ^(28)^.

### 2.5. Intervention

The BELL trial intervention involved moderate to high intensity hardstyle kettlebell training performed five days a week for 12 weeks. Face-to-face group-exercise was delivered three times weekly (Mon, Wed, Fri), and supplemented with prescribed home exercise which was performed twice-weekly (Tue/Thur). Due to COVID-19 restrictions, all training was performed at home during weeks 7-12 with training videos provided online, with the intervention period increased to 16 weeks due to final testing being delayed. Group classes were 45 minutes in duration and led by a certified hardstyle kettlebell instructor. Attendance to group sessions and compliance with home-exercise was recorded, together with internal and external training load volume. A modified CR10 scale was used for reporting sRPE ^(28)^. A training record which included exercise(s), number of sets, number of repetitions, and sRPE was maintained for analysis. Program design was based upon the principles and practices described by Tsatsouline ^(29)^ with exercises and delivery adjusted to account for individual limitations. The training period was preceded by a familiarisation week (2× 45-minute sessions) in which the participants were introduced to a standardised mobility drill and the foundational kettlebell exercises of a swing, clean, military press, goblet squat and unloaded Turkish get-up. Kettlebells for the different exercises and participants ranged from 4-80 kg.

All training sessions commenced with the standardised mobility routine which was used as a warm-up. During the first two weeks, participants were advised to work at a relatively low intensity (2-4/10: “easy” to “somewhat hard”) with a low volume training load to minimise the likelihood of experiencing delayed onset muscle soreness. From week three participants were encouraged to work up to a sRPE of 5-7/10 as tolerated (described as “hard” to “very hard”), with maximal effort (9-10/10) discouraged. Where technique was acceptable and RPE appeared to be <4/10, participants were encouraged to increase the kettlebell mass. Participants were able to self-select kettlebells and change any program variable within the group sessions. Supplementary home exercises, performed with an 8kg kettlebell (provided by the researcher) or bodyweight only, were prescribed with an achievable training load target with no upper limit on the total number of repetitions. Programming was based on i) physical capacity of the group, ii) participant feedback, iii) intent to offer variety, and iv) plan to progress skill, intensity, and training load volume throughout the intervention period. Due to COVID-19 restrictions, final testing was delayed by four weeks. During this time, participants performed 100 kettlebell swings daily. Training material and programming was conceived, delivered, coordinated, and analysed by the lead investigator (NM).

## 3. Results

Change in LS and FN BMD are shown in Table 2. Attendance rates for group sessions, compliance with prescribed home-exercise, total training load volume, and change in outcome measures pre-to post-training are shown in Table 3. Session training load volume and arbitrary units are shown in Fig. 1 and 2 for Sally and Peter, respectively.

**Table 2.** Change in LS and FN BMD pre- to post-training.

| Sally | 02/11/2017 (HOLOGIC)** |  |  | 21/01/2020 (HOLOGIC) |  | 11/08/2020 (NORLAND) |  |  |  |  |
| --- | --- | --- | --- | --- | --- | --- | --- | --- | --- | --- |
|  | Region | BMD (g/cm<sup>2</sup>) | T-score | BMD (g/cm<sup>2</sup>) | T-score | BMD (g/m<sup>2</sup>) | ΔBMD | BMC (g) | T-score | Change (%) |
|  | Fem neck | 0.579 | -2.4<sup>#</sup> | 0.596 | -2.3<sup>#</sup> | 0.6718 | 0.0758 | 2.276 | -2.21<sup>#</sup> | 12.7 |
|  | Troch |  |  |  |  | 0.5146 |  | 6.080 | -1.71<sup>#</sup> |  |
|  | Total sBMD | 669 mg/cm<sup>2</sup> | -2.2<sup>#</sup> | 669 mg/cm<sup>2</sup> | -2.2<sup>#</sup> | 680.3 mg/cm<sup>2</sup> | 0.0113 | 20889 mg | -2.15<sup>#</sup> | 1.6 |
|  | L2 - L4 | 0.736 | -3.1* | 0.748 | -3.0* | 0.7923 | 0.0443<sup>§</sup> | 33.11 | -2.31<sup>#</sup> | 5.9 |
|  | L2 |  |  |  |  | 0.8077 |  | 10.63 | -2.16<sup>#</sup> |  |
|  | L3 |  |  |  |  | 0.8334 |  | 11.05 | -2.31<sup>#</sup> |  |
|  | L4 |  |  |  |  | 0.7438 |  | 11.43 | -2.51* |  |
|  | Total sBMD |  |  |  |  | 852.6 mg/cm<sup>2</sup> |  | 35629 mg | -1.90<sup>#</sup> |  |
| Peter | 16/12/2019 (NORLAND) |  |  |  | 16/06/2020 (NORLAND) |  |  |  |  |  |
|  | Region | BMD (g/cm<sup>2</sup>) | BMC (g) | T-score | BMD (g/cm<sup>2</sup>) | ΔBMD | BMC (g) | T-score | Change (%) |  |
|  | Fem neck | 0.7195 | 3.907 | -3.11* | 0.7373 | 0.0178 | 3.981 | -2.97* | 2.5 |  |
|  | Troch | 0.7256 | 12.44 | -1.84<sup>#</sup> | 0.7308 | 0.0052 | 12.40 | -1.79<sup>#</sup> | 0.7 |  |
|  | Total sBMD | 880.4 mg/cm<sup>2</sup> | 33759 mg | -2.17<sup>#</sup> | 896.5 mg/cm<sup>2</sup> | 0.0161 | 34638 mg | -2.04<sup>#</sup> | 1.8 |  |
|  | L2 - L4 | 1.008 | 51.44 | -0.85 | 1.0680 | 0.060<sup>§</sup> | 55.47 | -0.52 | 6.0 |  |
|  | L2 | 1.026 | 15.93 | -0.78 | 1.0300 | 0.004 | 16.21 | -0.76 | 0.4 |  |
|  | L3 | 0.9974 | 17.83 | -0.96 | 1.0820 | 0.085<sup>§</sup> | 18.52 | -0.52 | 8.5 |  |
|  | L4 | 1.003 | 17.68 | -0.66 | 1.0870 | 0.084<sup>§</sup> | 20.74 | -0.24 | 8.4 |  |
|  | Total sBMD | 1085 mg/cm<sup>2</sup> | 55356 mg | -0.85 | 1149 mg/cm<sup>2</sup> | 0.064<sup>§</sup> | 59693 mg | -0.52 | 6.0 |  |

\*\* conducted before enrolling in the BELL trial, <sup>#</sup> osteopenia, \* osteoporosis, <sup>§</sup> > LSC
Troch; trochanteric, L1-L4; lumbar spine vertebrae 1-4, BMC; bone mineral content, sBMD; standardized bone mineral density

**Table 3.** Change in outcome measures, pre- to post-training.

|  |  | Sally |  |  |  | Peter |  |  |  |
| --- | --- | --- | --- | --- | --- | --- | --- | --- | --- |
| Attendance |  | 94.4 % |  |  |  | 94.4 % |  |  |  |
| Compliance |  | 87.5 % |  |  |  | 91.7 % |  |  |  |
| Training load (group sessions)<sup>o</sup> |  | 74,872 kg |  |  |  | 110,132 kg |  |  |  |
| Outcome measures | | Pre- | Post- | $\Delta$ | % | Pre- | Post- | $\Delta$ | % |
| Grip strength (kg) | *right* | 20 | 26 | 6<sup>e</sup> | 30.0 | 20 | 28 | 8<sup>e</sup> | 40.0 |
|  | *left* | 18 | 25 | 7<sup>e</sup> | 38.9 | 26 | 34 | 8<sup>e</sup> | 30.8 |
| 5x floor transfer time (s) |  | 31.1 | 21.4 | -9.7 | -31.2 | 41.2 | 38.8 | -2.4 | -5.8 |
| 6-min walk distance (m) |  | 677 | 740 | 63<sup>e</sup> | 9.3 | 584* |  |  |  |

<sup>e</sup> > minimum clinically important difference (grip = 5kg, 6MWD = 50m)
<sup>o</sup> excludes prescribed home-training sessions Tue/Thur
\* Peter did not complete the final 6-min walk test.

**Figure 1.**
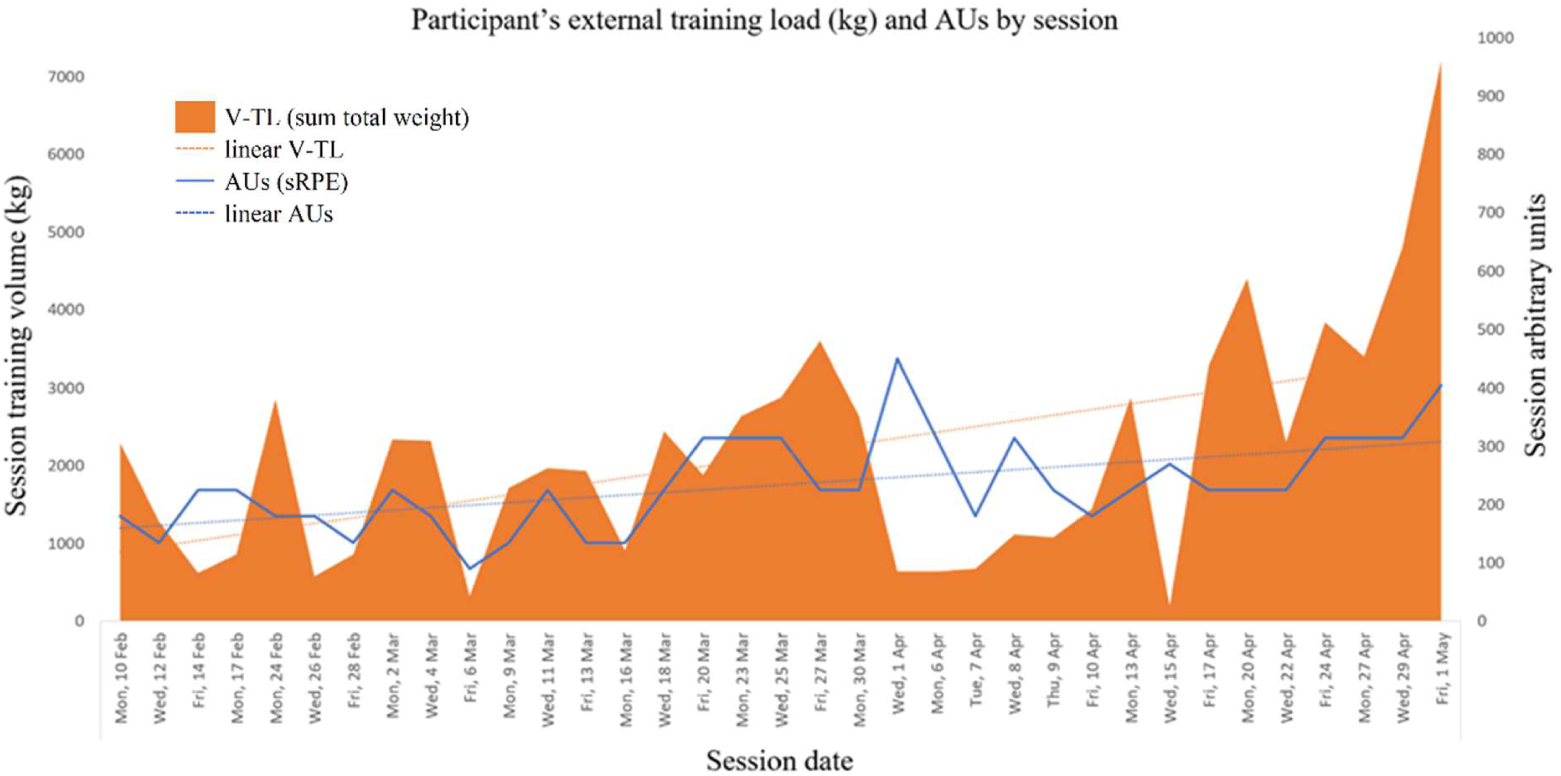
12-week training load - *Sally*.

**Figure 2.**
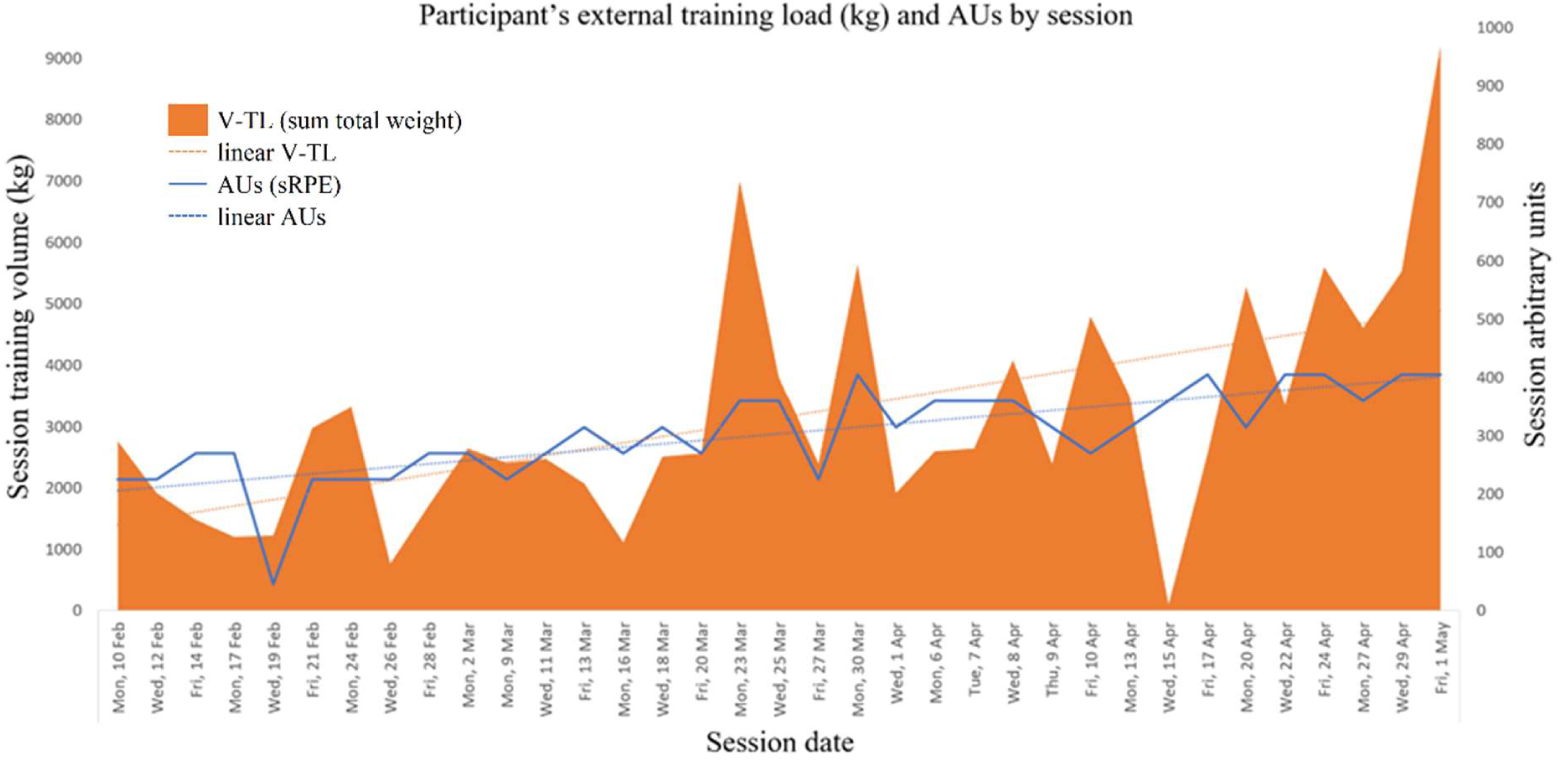
12-week training load - *Peter*.

Sally and Peter’s change in training load are representative of the BELL trial cohort. Internal and external training load volume depict a model of undulating periodisation, which steadily increased throughout the intervention period. The final two weeks training volume was prescribed, to enable all participants to attain a training load ‘personal best’ on the final day of the trial. Training at home with only limited kettlebells, Sally and Peter both recorded a “very hard” sRPE of 9/10 Friday 1 May.

Sally missed four consecutive training sessions due to a bereavement, and one further home training session. Peter missed four consecutive training sessions due to sickness. Least significant change in BMD was calculated to be 0.022g/cm^2^ for Sally, and 0.033g/cm^2^ for Peter. For Sally, an increase in BMD of 0.017 g/cm^2^ at the FN and 0.012 g/cm^2^ at the LS between 2 November 2017 and 21 January 2020 (before the BELL trial intervention) was below the LSC calculated from her BMI. Between 21 January 2020 and 11 August 2020 (following the intervention), a BMD increase of 0.0443 g/cm^2^ at the LS was more than double her LSC (2.01×) and sufficient to advance her status from osteoporosis to osteopenia. In addition, Sally’s FN BMD increased by 12.7%. Between 16 December 2019 and 16 June 2020, Peter’s total lumbar spine BMD increased by 0.064 g/cm^2^, almost double his LSC (1.94×). Peter’s femoral neck BMD also increased by 2.5%.

## 4. Discussion

This case study is the first to show clinically significant increases in bone mass in older adults with osteoporosis, following 16-weeks of hardstyle kettlebell training. These results challenge the view that a combination of high intensity, and impact loads, are necessary for an osteogenic exercise effect ^(4)^. Change in outcome measures and training load volume for both subjects in this study are representative of the results from the BELL trial, which show large improvements in grip strength, and significant increases in cardiovascular and functional capacity. Although these data are not definitive evidence of a causal relationship between kettlebell training and increased bone mass in older adults with osteoporosis, these findings warrant further investigation in sufficiently powered, randomised controlled trials.

The predictive value of low BMD for hip fracture is comparable to that of high blood pressure for stroke ^(6)^. Bone metabolism deteriorates with advancing age with insufficiently active older adults typically losing 1-3% BMD every year ^(30)^. Rates of bone loss can be exacerbated by inactivity, disuse, medical conditions, and deficiencies in calcium and vitamin D ^(31)^. Although osteoporosis is influenced by epigenetic and environmental factors ^(18)^, and mediated by factors other than mechanical loading ^(32)^, physical activity and exercise are a major extrinsic factor responsible for the maintenance and promotion of bone mass ^(33, 34)^. Being physically active is recommended for all adults ^(35)^ however, systematic reviews remain cautious of overstating the preventative potential of physical activity for preventing osteoporosis ^(36)^. Movement behaviour, specifically moderate-to-vigorous physical activity, is significantly associated with increased bone mass ^(37)^ and reduced risk of hip fracture, of 45% and 38% in men and women respectively ^(38)^.

The most recent and comprehensive review and meta-analysis to date, of exercise intensity on bone in postmenopausal women, highlights the notable absence of trials using high intensity (>80% 1RM) training ^(39, 40)^. Only six studies met the inclusion criteria of ‘high intensity’ of which only the LIFTMOR trial^(23)^ reported a statistically significant increase in LS BMD. Consistent with previous meta-analyses ^(14, 24, 41-43)^ the within-group mean increase in LS BMD in the LIFTMOR trial, was 0.023g/cm^2^ (2.9 ± 3.1%) with no significant change in FN BMD. A change of that magnitude at the lumbar spine is 0.001g/cm^2^ above the LSC for the underweight female in the current study, and 0.01g/cm^2^ below the LSC for the male whose BMI was only slightly above ‘normal’. Subsequent iterations of the HiPRT protocol used in the LIFTMOR trial have been underpowered to detect a change in bone mass ^(44)^, or resulted in a mean change of smaller magnitude (0.016g/cm^2^) ^(45)^. In the meta-analysis by Kistler-Fischbacher et al ^(40)^ the authors reported, 1) no evidence that high intensity training was more effective at the femoral neck, 2) no significant difference in effect size between moderate and high intensity training, 3) no significant effects of impact-only exercises regardless of intensity, 4) no benefit of resistance and impact training combined compared to resistance-only training, and 5) no association between resistance training intensity (measured as a percentage of 1RM) and change in LS and FN BMD.

Results from other trials and meta-analyses have been mixed. Single trials have reported no significant difference between training frequencies ^(46)^, no significant difference between resistance training and aerobic training performed over two years ^(47)^, and no significant change following three years of resistance training ^(48)^. Outcomes have also been shown to become non-significant when non-randomised trails, are excluded from analysis ^(16)^, likely due to the confounding associated with non-randomised trials which can almost double reported treatment effects. Other meta-analyses have reported that only studies with combined aerobic and strength training programs, result in improvements likely to reach a least significant change ^(41)^. These are consistent with other data indicating an overall treatment effect <1% ^(16, 49)^ irrespective of concurrent HRT, bisphosphonate or calcium supplementation ^(24)^. Aerobic-only and impact-only programs are largely ineffective when used in isolation, but programs which combine aerobic/impact exercise with resistance training are more likely to result in a weighted mean difference of >0.02g/cm^2 (16)^. The findings of this study suggest there are other mediators of an osteogenic effect besides impact and high intensity loads.

The BELL and LIFTMOR trials both reported improvements in functional outcomes and bone health. The LIFTMOR trial utilised a barbell deadlift, overhead press, and back squat performed at 80 to 85% 1RM, with drop landings from a chin-up bar. Participants in the LIFTMOR trial completed a total of 75 repetitions per session for 70 sessions over an 8-month training period, compared to the 60 sessions in 3 months during the BELL trial. Mean training load was not reported in the LIFTMOR trial but given the maximum number of weighted repetitions per session (75), it is likely to have been less that the 2805 ± 251 kg per session reported in the BELL trial. Rating of perceived exertion was high in the BELL trial with all participants recording an sRPE value of 8-10/10 (“very hard” to “maximal”) on the final day, but training involved no impact loading and only three sessions in which participants may have performed kettlebell deadlifts with loads >85% 1RM. For the last 10 weeks of the training period, participants only had access to three kettlebells to train at home; Sally had 8, 12 and 24 kg, Peter had two 8s and 22 kg.

A 12-month trial of resistance training with a much larger external training load than the BELL trial, found no significant relationship between training load and BMD ^(50)^, casting doubt over the influence of training load volume as a mediator. A positive relationship between BMD and training volume has been reported in men, but this may be limited to impact loads only ^(51)^, thus mediators of the osteogenic effect observed in the current case study remain to be determined. Some evidence does indicate that high velocity resistance exercises may have a superior effect on FN and LS BMD than performing the same movements more slowly ^(52)^. In support of this view, net peak ground reaction force in older adults, was greater during a two-handed hardstyle swing with an 8 kg kettlebell, than a kettlebell deadlift with 32 kg ^(53)^, and data from a pilot study of low load power training in older adults, shows this combination of high velocity and light load may be sufficient for osteogenesis to occur ^(26)^. Given the ballistic nature of the hardstyle kettlebell exercises used in the BELL trial, the high velocity of kettlebell swings compared to standard resistance training exercises may help explain the results of this study.

Most studies measuring the effects of interventions for osteoporosis, rely on DXA-derived BMD as a surrogate measure of bone strength, and a proxy for primary clinical outcomes, namely fractures ^(8, 13)^. This can be somewhat problematic because bone strength is influenced by factors other than BMD, which the planar nature of DXA is not ideally suited to measure, such as bone size, shape, and structure. Furthermore, small changes in cross-sectional area can improve strength, independent of BMD ^(13)^. Significant increases in cortical volume and thickness, without corresponding increases in trabecular BMC and total volume, as reported in the LIFTMOR trials ^(23, 44)^, might be explained, in part at least, by improved edge detection resulting in a larger area of bone being reported ^(54)^ or redistribution of bone minerals ^(55)^.

Indices of bone strength as outcomes have been limited ^(39)^, so the effects of exercise on the properties which improve bone strength, are not currently well known, and definitive conclusions cannot yet be made, regarding the effectiveness of exercise for improving bone strength and reducing fracture risk ^(6, 8, 13)^. Increased bone mass is not confirmation of an improvement in bone quality or strength ^(2)^, therefore the effectiveness of an intervention should be measured by its influence on reducing incidence of fracture, rather than change in BMD. Unfortunately, evidence is conflicting whether exercise reduces fracture risk ^(56)^, and it is presently not feasible to conduct a definitive trial to determine that ^(4)^.

Underscoring the potential prophylactic nature of exercise, clinical guidelines recommend that exercise should be lifelong, and include regular weight-bearing and high impact activities to promote muscle strength, muscle power, and maintenance of BMD ^(6, 14, 18)^. Resistance exercise is recommended for older adults because it attenuates bone loss and enhances physical function ^(57)^. Regular and consistent exercise which provides sufficient loading is especially important for bone mass and geometry ^(8)^, yet 50 % of individuals with osteoporosis who start a new exercise program have been reported to discontinue within 6 months ^(20)^. Programs are needed which have high rates of engagement long-term, and which provide older adults with the motivation, self-efficacy, and skills, to be able to perform at least some of their exercise independently.

A case study of a 70-year-old postmenopausal women, reported a remarkable 24.3% increase in bone mass (0.196g/cm^2^) at the femoral neck and 29.7% (0.17g/cm^2^) at the lumbar spine ^(22)^, following an extensive 12-month program of resistance training. A limitation reported in the BELL trial was that the group format was not ideally suited to the individuals with high or low functional capacity ^(21)^. The remarkable effects reported by Aquino et al. ^(22)^ may be a product of the N-of-1 design, which allows the exercise program to be uniquely tailored to meet the needs and physical capacity of the individual. This is encouraging for healthcare providers training with older adults 1-on-1, but potentially challenging for those designing group-based programs. It is feasible that the rates of increase in bone mass observed in the present study, could be maintained with ongoing training, and may achieve a similar magnitude of effect to that reported by Aquino et al ^(22)^ with similar a training duration.

Due to their fear of fracture, older adults diagnosed with osteoporosis are often overly cautious and risk-averse towards engaging in impact and resistance exercise ^(7)^. This fear and anxiety can be compounded by inappropriate messages from healthcare providers where uncertainty exists around what is appropriate and safe ^(6)^. Inadequate and inconsistent advice can cause confusion and may have a negative effect on the person’s social and psychological health, wellbeing, and self-efficacy ^(8)^. Promoting the benefits of exercise can be framed positively. For most older adults, engaging in activities which promote muscle and bone strength, is safe and will help improve or at least maintain function ^(8)^, and exercise does not have to be vigorous to be beneficial. Providing reassurance to older adults with low bone mass, and encouraging physical activities which promote bone health, should be a priority for healthcare providers ^(6)^.

Results from a qualitative study of the BELL trial, revealed the effectiveness and perceived health benefits of older adults participating in group kettlebell training with similar age peers ^(21)^. Participants reported being more physically active, having increased self-confidence, an improved physical capacity, and enhanced mental health following the trial. Age, training status, sex, and the presence of comorbid health conditions, such as osteoarthritis and persistent pain, did not negatively impact engagement or participation in the training. Attendance was >90% with some participants, including Sally, still independently training together more than 18 months after the BELL trial intervention ended.

Regular exercise can effectively reduce a myriad of diseases associated with ageing. Exercise has been described as a true geroprotector, with even modest amounts of exercise capable of improving quality of life and increasing lifespan ^(58)^. It has been suggested that 12 weeks of exercise is insufficient to detect physiological change in bone, as the remodelling cycle typically lasts 3-8 months ^(19, 59)^, however, data from this and other studies appears to dispute that ^(60)^. Increases in total hip BMD of 6% have also been reported from 16 weeks of resistance training in elderly women with decreased muscle strength ^(61)^.

Results from the BELL trial demonstrated that hardstyle kettlebell training can improve functional measures of healthy ageing. Kettlebell training can be performed safely and independently by older adults, and when performed in a group setting, can be very rewarding, even at high rates of perceived exertion. Improvements in lean mass from exercise is protective against bone loss and risk of vertebral fractures, as anabolic signalling in the bone-muscle unit works both ways: muscle influences bone and vice versa ^(31, 34)^. The mean increase of 0.65 kg in axial skeletal muscle mass reported in the BELL trial ^(1)^, adds further support for kettlebell training being an effective mode of resistance training, to promote healthy ageing in older adults. Results of the present study suggests that kettlebell training might also be an effective intervention to promote bone health in older adults with low or very low BMD.

These case findings are not without limitations. Firstly, Hologic and Norland machines have different methods of deriving energy beams, different calibration techniques, and different methods for determining the regions of interest for analysis. Although comprehensive evaluation of the DXA-measured BMD values obtained using Hologic and Norland densitometers have been found to be virtually the same ^(62)^, with in-vivo comparison of spinal measurements highly correlated (BMD: r = 0.990, SEE = 0.028 g/cm^2^) and comparable in terms of precision and accuracy ^(63)^, the degree of measurement error inherent between Sally’s reports cannot be determined. Up to approximately 50% of the 0.064g/cm^2^ change in BMD observed at Peter’s lumbar spine, could be an effect of the Denosumab he received in the weeks preceding commencement of training ^(64)^.

## 5. Conclusion

Results of this study suggest that 16-weeks of high-volume moderate to high-intensity hardstyle kettlebell training performed 5 days a week, can improve bone mineral density in older adults with osteoporosis, with magnitude of change sufficient to advance osteoporosis to osteopenia. Kettlebell training may therefore be a viable mode of exercise for improving bone health and reducing fracture risk in older adults with low and very low BMD. For the two participants in the current study, the nature of the relationship between the training intervention and the observed increase in BMD remains unclear. However, taken together with the clinically significant improvements in other outcome measures, high rates of engagement, and positive participant experiences, we suggest further research in randomised controlled trial is warranted.

### Plain language summary

Clinically significant increases in bone mass were observed in two older adults with osteoporosis, following moderate to high intensity hardstyle kettlebell training performed five days a week for 16 weeks, sufficient to advance osteopenia to osteoporosis.

## Supporting information

CARE checklist

## Data Availability

Data will be made available in accordance with Bond University Research Data Management and Sharing Policy (TLR 5.12, Sections 9.3, 10.2. 10.5, 10.8). Research data and primary materials will be made openly available for use by other researchers and interested persons for further research after a reasonable period following completion of the research.

## Acknowledgements

The authors would like to thank the case subjects for making their DXA reports available for publication, and Dr Ben Schram for reviewing the final manuscript.

## Contributions

NM curated and analysed the data, interpreted the results, conducted the formal analysis, and wrote the original draft. JK and WH supported with ongoing consultation, reviewed, and provided revisions to the manuscript. All authors read and approved the final manuscript.

## Ethics declarations

### Ethics approval and consent to participate

The study was approved by the Bond University Human Research Ethics Committee (NM03279) which was subsumed within a larger clinical trial. The BELL trial was pre-registered on the Australian New Zealand Clinical Trials Registry (ACTRN12619001177145). Permission to reproduce BMD reports was granted by both subjects.

### Consent for publication

Permission to reproduce BMD reports was granted by both subjects.

## Rights and permissions

This article is distributed under the terms of the Creative Commons Attribution 4.0 International License (http://creativecommons.org/licenses/by/4.0/), which permits unrestricted use, distribution, and reproduction in any medium, provided you give appropriate credit to the original author(s) and the source, provide a link to the Creative Commons license, and indicate if changes were made. The Creative Commons Public Domain Dedication waiver (http://creativecommons.org/publicdomain/zero/1.0/) applies to the data made available in this article, unless otherwise stated.

## Supplementary data

- The BELL trial CARE (CAse REport) checklist

