## Supplementary material for "Effect of kettlebell training on bone mineral density in two older adults with osteoporosis: a multiple-case study from the BELL trial": CARE checklist

Australian New Zealand Clinical Trials Registry (ID: ACTRN12619001177145).

**CARE (CAse REport) guidelines**

| Item name | Item | Brief description | Addressed on page number/ heading |
|---|---|---|---|
| Title | 1 | The words "case report" (or "case study") should appear in the title along with phenomenon of greatest interest (e.g., symptom, diagnosis, test, intervention) | p1 |
| Key words | 2 | The key elements of this case in 2-5 words | p1 |
| Abstract | 3 | a) Introduction - What does this case add?<br>b) Case Presentation:<br>• The main symptoms of the patient<br>• The main clinical findings<br>• The main diagnoses and interventions<br>• The main outcomes<br>c) Conclusion - What were the main "take-away" lessons from this case? | p1 |
| Introduction | 4 | Brief background summary of this case referencing the relevant medical literature | p1-2 |
| Participant information | 5 | a) Demographic information (e.g., age, gender, ethnicity, occupation)<br>b) Main symptoms of the patient (his or her chief complaints)<br>c) Medical, family, and psychosocial history - including diet, lifestyle, and genetic information whenever possible, and details about relevant comorbidities including past interventions and their outcomes | p2<br>n/a<br>p2 |
| Clinical findings | 6 | Describe the relevant physical examination (PE) findings | p5 |
| Timeline | 7 | Depict important dates and times in this case (table or figure) | p4-5 |
| Diagnostic assessment | 8 | a) Diagnostic methods (e.g., PE, laboratory testing, imaging, questionnaires)<br>b) Diagnostic challenges (e.g., financial, language/cultural)<br>c) Diagnostic reasoning including other diagnoses considered<br>d) Prognostic characteristics (e.g., staging) where applicable | p5<br>n/a<br>n/a<br>n/a |
| Therapeutic intervention | 9 | a) Types of intervention (e.g., pharmacologic, surgical, preventive, self-care)<br>• Administration of intervention (e.g., dosage, strength, duration)<br>• Changes in intervention (with rationale) | p3-4 |
| Follow-up and outcomes | 10 | a) Summarize the clinical course of all follow-up visits including<br>• Clinician and patient-assessed outcomes<br>• Important follow-up test results (positive or negative)<br>• Intervention adherence and tolerability (and how this was assessed)<br>• Adverse and unanticipated events | p5 |

| Item name | Item | Brief description | Addressed on page number/ heading |
|---|---|---|---|
| Discussion | 11 | a) The strengths and limitations of the management of this case<br>b) The relevant medical literature<br>c) The rationale for conclusions (including assessments of cause and effect) d) The main ‘‘take-away’’ lessons of this case report | p8<br>p6-8<br>p7-8 |
| Participant perspective | 12 | The patient should share his or her perspective or experience whenever possible | n/a |
| Informed consent | 13 | Did the patient give informed consent? Please provide if requested | Yes, p2 |

*Gagnier, Joel J., et al. "The CARE guidelines: consensus-based clinical case report guideline development." Journal of clinical epidemiology 67.1 (2014): 46-51.*
